# Fully Automated Multi-Chamber Strain Profiling in Consecutive Patients Undergoing Transthoracic Echocardiography: Feasibility, Functional Clustering, and Prognostic Implications

**DOI:** 10.64898/2026.10.20.26365880

**Authors:** Su A Noh, Jiesuck Park, Yeonyee E. Yoon, Jaeik Jeon, Yeonggul Jang, Seung-Ah Lee, Minjung Bak, Hong-Mi Choi, In-Chang Hwang, Goo-Yeong Cho

## Abstract

**Aims:** Myocardial strain enables sensitive assessment of cardiac function, but its clinical application has largely focused on individual chambers or selected patient populations. We evaluated the feasibility and clinical relevance of fully automated multi-chamber strain assessment in consecutive patients undergoing routine transthoracic echocardiography (TTE).

**Methods and results:** This retrospective study included 8,350 consecutive adults undergoing TTE. A hospital-wide artificial intelligence (AI)–based system automatically measured left ventricular (LV) global longitudinal strain (LVGLS), left atrial (LA) reservoir strain (LASr), and right ventricular (RV) free-wall strain (RVFWS). AI-derived LVGLS was available from at least one apical view in 97.1% of patients and from all three views in 94.8%; LASr and RVFWS were available in 94.1% and 94.5%, respectively, resulting in complete multi-chamber strain assessment in 90.3%. Strain measures showed modest correlations with conventional chamber-specific echocardiographic parameters and differential associations with clinical outcomes. Gaussian mixture modeling identified three distinct clusters characterized by preserved multi-chamber function, RV-dominant dysfunction, and pan-chamber dysfunction. Compared with the preserved cluster, pan-chamber dysfunction was associated with the highest risk of all-cause death or heart failure hospitalization (adjusted hazard ratio, 3.44; 95% CI, 2.43–4.88; P<0.001). Prognostic associations remained evident in patients with LV ejection fraction (LVEF) ≥50%, and cluster assignment provided incremental prognostic information beyond clinical factors, LVEF, and RV fractional area change.

**Conclusions:** Fully automated multi-chamber strain assessment was feasible in a large consecutive TTE cohort. Integrating strain across cardiac chambers may provide complementary functional characterization and identify clinically relevant clusters with prognostic information beyond conventional assessment.

**Graphical Abstract:** 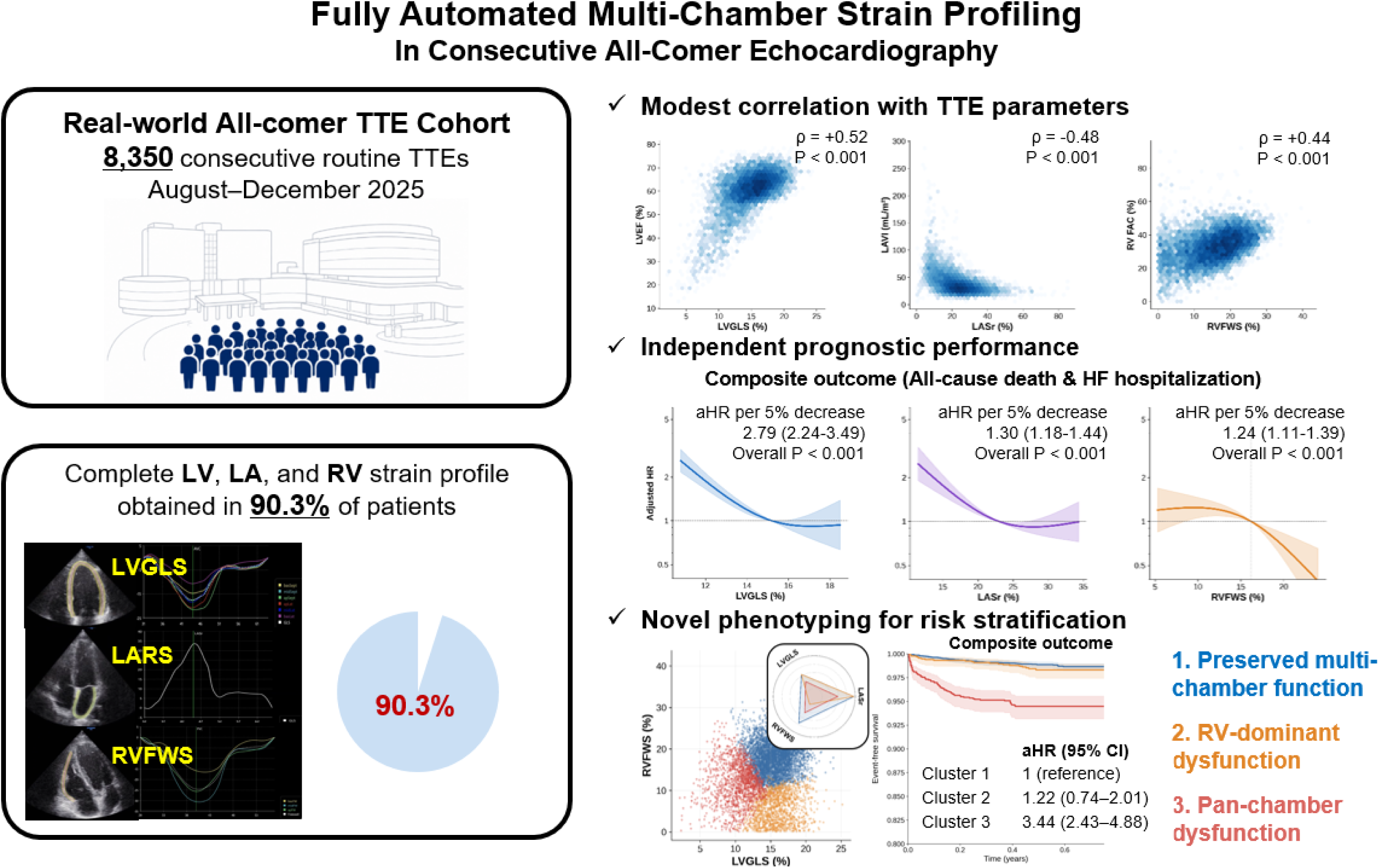

## INTRODUCTION

Transthoracic echocardiography (TTE) is central to assessing cardiac structure and function across a broad spectrum of clinical conditions. In most echocardiographic examinations, cardiac chamber structure and function are assessed using conventional chamber-specific measures, such as left ventricular (LV) ejection fraction (LVEF), left atrial (LA) volume, and right ventricular (RV) fractional area change (FAC).^1^ Although these measures provide important diagnostic and prognostic information, they may be influenced by loading conditions, geometric assumptions, and measurement variability and may not fully capture subtle myocardial dysfunction.^2^ Myocardial deformation imaging provides a sensitive and quantitative assessment of cardiac function beyond conventional echocardiographic measures. LV global longitudinal strain (LVGLS) can identify systolic impairment that may not be apparent from LVEF alone, whereas LA reservoir strain (LASr) reflects atrial compliance, myocardial function, and the cumulative effects of LV filling pressure.^2,3^ RV free-wall strain (RVFWS) provides complementary information regarding RV systolic function beyond conventional measures.^2,3^ Accordingly, strain imaging has emerged as a valuable tool for characterizing myocardial dysfunction in a variety of cardiovascular conditions.

Despite the growing clinical evidence supporting myocardial strain, most clinical applications and previous studies have evaluated strain within a single cardiac chamber or focused on selected disease populations.^4–6^ However, abnormalities in LV, LA, and RV function frequently coexist and may reflect different components of a shared hemodynamic and pathophysiologic process. Simultaneous assessment of strain across multiple chambers may therefore provide a more comprehensive representation of cardiac dysfunction than assessing any single strain measure alone. Furthermore, combined LV, LA, and RV strain patterns may reveal clinically distinct clusters of cardiac dysfunction not captured by isolated chamber assessment. In this study, we evaluated the feasibility and clinical relevance of a hospital-wide artificial intelligence (AI)–based system integrated into the TTE workflows for fully automated measurement of LV, LA, and RV strain in a real-world cohort of consecutive patients. We further aimed to characterize multi-chamber strain patterns across clinical settings, assess their associations with conventional TTE parameters, identify distinct multi-chamber strain clusters, and determine their association with clinical outcomes.

## METHODS

### Study Design and Population

This retrospective observational study included 8,350 consecutive patients who underwent TTE at the adult echocardiography laboratory of Seoul National University Bundang Hospital between August and December 2025. An AI-based analysis system had been integrated into the hospital echocardiographic infrastructure before the study period and operated as part of the clinical TTE workflow. All eligible TTE examinations were automatically processed for LV, LA, and RV strain analysis, regardless of the TTE indication or clinical care setting.

We included the entire cohort in the feasibility analysis. Then, we restricted analyses of multi-chamber strain patterns, clustering, and clinical outcomes to patients with successful AI-derived strain measurements in all three cardiac chambers (**Supplemental Method 1)**. The Institutional Review Board (B-2607-1054-109) approved the study protocol and waived informed consent because the study used retrospective, anonymized data.

### Automated Analysis Workflow and Multi-Chamber Strain Analysis

This study used the AI-based echocardiographic analysis system Sonix Health (Ontact Health Co., Ltd., Republic of Korea). After each examination, TTE images were automatically transferred to a dedicated analysis server without a separate referral, manual image transfer, or operator initiation. The system then automatically identified relevant echocardiographic views and performed cardiac chamber segmentation and motion analysis to generate conventional echocardiographic measurements as well as myocardial strain parameters.^7–12^

LVGLS, LASr, and RVFWS were generated through automated myocardial motion tracking without manual view selection, contour adjustment, tracking correction, or parameter entry.^9,11,12,13^ When no suitable view was available, or an image did not meet the predefined requirements for segmentation, measurement, or motion tracking, the corresponding parameter was not generated. No additional image acquisition was performed, and unsuccessful analyses were not manually corrected or retrospectively reanalyzed. Details of the automated analysis methods are provided in **Supplemental Methods 2**. AI-derived strain results were stored and displayed in the hospital electronic medical record (EMR) system, as shown in **Figure 1**.

**Figure 1.**
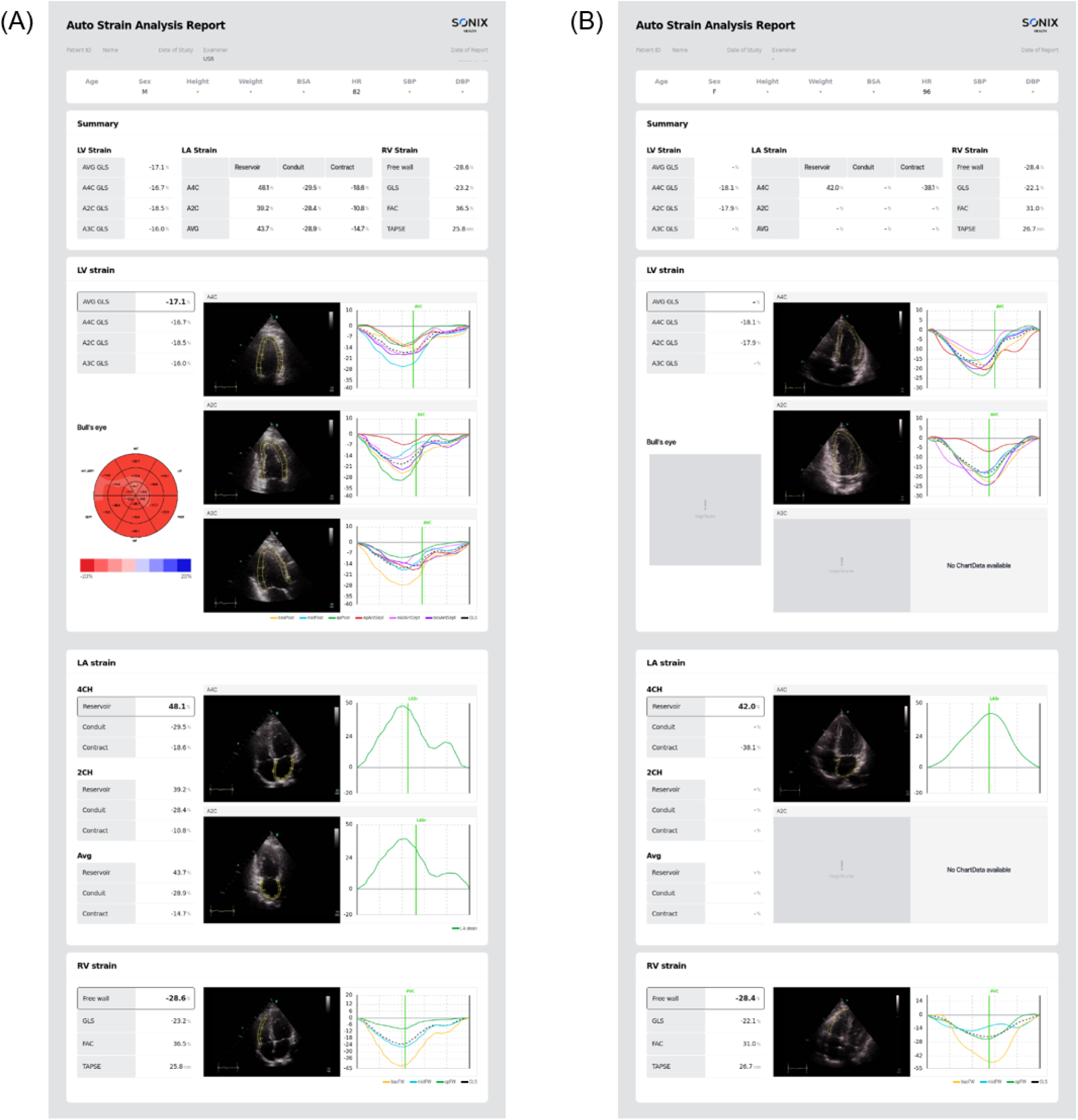
Clinical Display of Automated Multi-Chamber Strain Analysis. Representative EMR displays of (A) successful and (B) unsuccessful automated multi-chamber strain analysis.

For study analyses, AI-derived results were linked to clinical and conventional echocardiographic data using anonymized study identifiers. Chamber-specific feasibility was defined by generation of a valid strain value. Successful multi-chamber strain analysis was defined as the simultaneous availability of complete three-view LVGLS, apical 4-chamber view (A4C)-derived LASr, and RVFWS measurements from the same index TTE examination. Complete three-view LVGLS measurement required successful analysis of all three apical views, while A4C-derived LASr was selected in accordance with current consensus recommendations.^3^ A2C-derived LASr was assessed only for view-specific technical feasibility. All strain measures were expressed as absolute values, with higher values indicating better myocardial deformation.

### Data Collection and Conventional Echocardiographic Assessment

Baseline demographic information and clinical data were collected from the EMR. TTE was performed using commercially available ultrasound systems and standard echocardiographic measurements were obtained in accordance with contemporary guideline recommendations as part of routine clinical care.^2^ Conventional echocardiographic parameters were preferentially obtained from clinical TTE reports, with AI-derived measurements substituted when unavailable. RV FAC was predominantly obtained from automated analysis because it was not routinely measured clinically. **Supplemental Methods 3** and **4** provide details on measurement availability, AI substitution, and manual–AI agreement.

### Clinical Outcomes

The primary outcome was a composite of all-cause death and heart failure (HF) hospitalization, with each component analyzed separately as secondary outcomes. Outcomes were ascertained from the EMR; all-cause death was identified from death certificates, and HF hospitalization was defined as hospitalization with a primary discharge diagnosis of HF. Follow-up continued from the index TTE date until the first occurrence of the outcome or censoring on May 14, 2026.

### Statistical Analysis

Continuous variables are expressed as medians with interquartile ranges (IQR) and categorical variables as counts with percentages. Between-cluster differences were assessed using the Kruskal–Wallis and chi-square tests, as appropriate. Spearman correlation coefficients were used to assess associations between strain and conventional TTE parameters.

Prognostic associations of AI-derived strain measures were evaluated using multivariable Cox proportional hazards models adjusted for age, sex, and modified Charlson Comorbidity Index (mCCI), including myocardial infarction, heart failure, stroke, chronic pulmonary disease, liver disease, diabetes mellitus, end-stage renal disease, and malignancy. Continuous associations were assessed using restricted cubic splines with knots at the 10th, 50th, and 90th percentiles and the cohort median as reference; overall and nonlinear associations were tested using likelihood-ratio tests. Hazard ratios per 5-percentage-point decrease in strain were estimated from linear Cox models with identical adjustment.

Gaussian mixture modeling (GMM) with a tied covariance structure was used to identify multi-chamber strain clusters based on standardized absolute LVGLS, LASr, and RVFWS. The number of clusters was selected considering the Bayesian information criterion, integrated completed likelihood, silhouette analysis, bootstrap-adjusted Rand index, parsimony, and clinical interpretability. Associations between cluster assignment and clinical outcomes were evaluated using Kaplan–Meier analysis and multivariable Cox proportional hazards models, adjusted for the same covariates.

Incremental prognostic value of cluster assignment was assessed by comparing two nested Cox proportional hazards models: Model 1 included age, sex, mCCI, LVEF, and RV FAC, whereas Model 2 additionally included cluster assignment. Overall model fit was summarized using global likelihood-ratio chi-square statistics, and the incremental contribution of cluster assignment was assessed using a likelihood-ratio test comparing Model 2 with Model 1. Subgroup analyses of the associations between cluster assignment and clinical outcomes were performed in patients with LVEF ≥50% using the same multivariable Cox modeling strategy.

All analyses were performed using Python (version 3.12) with the pandas, NumPy, SciPy, scikit-learn, and lifelines libraries. A two-sided P <0.05 was considered statistically significant.

## RESULTS

### Study Population and Feasibility

A total of 8,350 consecutive patients underwent TTE during the study period (**Supplemental Method 1**). AI-derived LVGLS was available from at least one apical view in 8,109 patients (97.1%), while complete three-view LVGLS using the A4C, A2C, and A3C was available in 7,915 patients (94.8%) (**Table 1**). For LA strain analysis, LASr was available from the A4C in 7,858 patients (94.1%). RVFWS was successfully generated in 7,887 patients (94.5%). Overall, complete multi-chamber strain data, defined as the simultaneous availability of three-view LVGLS, A4C-derived LASr, and RVFWS, were available in 7,539 patients (90.3%), who constituted the final analytic cohort. The remaining 811 patients (9.7%) lacked at least one required chamber-specific strain measurement. **Figure 1** shows representative examples of examinations with successful and unsuccessful automated multi-chamber strain analysis.

**Table 1.** Feasibility of Fully Automated Chamber-Specific and Multi-Chamber Strain Analysis.

| Strain | View | N | % |
| --- | --- | --- | --- |
| LVGLS | **All three views (A4C, A2C, A3C)\*** | 7,915 | 94.8 |
|  | Any analyzable view | 8,109 | 97.1 |
|  | A4C | 8,071 | 96.7 |
|  | A2C | 8,030 | 96.2 |
|  | A3C | 8,013 | 96.0 |
| LASr | Any analyzable view | 8,059 | 96.5 |
|  | **A4C†** | 7,858 | 94.1 |
|  | A2C | 7,720 | 92.5 |
| RVFWS | **Prespecified RV apical-view hierarchy** | 7,887 | 94.5 |
| Multi-chamber strain analysis‡ | **Complete 3-view LVGLS + A4C LASr + RVFWS** | 7,539 | 90.3 |

\* LVGLS was calculated as the average of valid longitudinal strain measurements obtained from all apical views (A4C, A2C, and A3C) and was used for subsequent analyses.
† A4C-derived LASr was used for subsequent analyses; A2C-only measurements were considered only in the technical feasibility assessment.
‡ Multi-chamber strain analysis was defined as the simultaneous availability of 3-view LVGLS, A4C-derived LASr, and RVFWS.
Abbreviations: LVGLS, left ventricular global longitudinal strain; LASr, left atrial reservoir strain; RVFWS, right ventricular free-wall strain; A4C, apical four-chamber; A2C, apical two-chamber; A3C, apical three-chamber.

### Clinical and Echocardiographic Characteristics According to Analysis Feasibility

Baseline characteristics according to the feasibility of complete multi-chamber strain analysis are summarized in **Table 2**. Compared with patients with successful analysis, those with unsuccessful analysis had a higher prevalence of heart failure, atrial fibrillation, and a modestly higher mCCI. They were also more frequently treated with mineralocorticoid receptor antagonists and loop diuretics. LVEF was similar between the two groups, whereas unsuccessful analysis was associated with higher E/e′, lower e′ velocity, slightly higher peak tricuspid regurgitation velocity (TR V_max_), and lower RV FAC.

**Table 2.** Baseline Clinical and Echocardiographic Characteristics According to Feasibility of Multi-Chamber Strain Analysis.

| **Variable** | **Overall**<br>**(N = 8,350)** | **Successful**<br>**multi-chamber strain**<br>**analysis**<br>**(N = 7,539)** | **Unsuccessful**<br>**multi-chamber strain**<br>**analysis**<br>**(N = 811)** | **SMD** | **P value** |
| --- | --- | --- | --- | --- | --- |
| **Demographics** |  |  |  |  |  |
| Age, years | 69.0 (60.0–77.0) | 69.0 (60.0–77.0) | 71.0 (60.0–79.0) | 0.09 | <0.001 |
| Male | 4798 (57.5) | 4366 (57.9) | 432 (53.3) | 0.09 | 0.012 |
| **Comorbidities** |  |  |  |  |  |
| Hypertension | 1854 (22.2) | 1678 (22.3) | 176 (21.7) | 0.01 | 0.751 |
| Diabetes mellitus | 444 (5.3) | 392 (5.2) | 52 (6.4) | 0.05 | 0.168 |
| Heart failure | 1277 (15.3) | 1102 (14.6) | 175 (21.6) | 0.18 | <0.001 |
| Myocardial infarction | 662 (7.9) | 605 (8.0) | 57 (7.0) | 0.04 | 0.353 |
| Atrial fibrillation | 1394 (16.7) | 1211 (16.1) | 183 (22.6) | 0.17 | <0.001 |
| COPD | 251 (3.0) | 211 (2.8) | 40 (4.9) | 0.11 | 0.001 |
| Stroke | 1002 (12.0) | 881 (11.7) | 121 (14.9) | 0.10 | 0.008 |
| ESRD | 239 (2.9) | 215 (2.9) | 24 (3.0) | 0.01 | 0.949 |
| Liver cirrhosis | 111 (1.3) | 98 (1.3) | 13 (1.6) | 0.03 | 0.579 |
| Malignancy | 1841 (22.0) | 1659 (22.0) | 182 (22.4) | 0.01 | 0.810 |
| mCCI | 1.0 (0.0–2.0) | 1.0 (0.0–2.0) | 1.0 (0.0–2.0) | 0.12 | 0.001 |
| 0 | 4097 (49.1) | 3742 (49.6) | 355 (43.8) |  |  |
| 1–2 | 3420 (41.0) | 3072 (40.7) | 348 (42.9) |  |  |
| 3–4 | 766 (9.2) | 667 (8.8) | 99 (12.2) |  |  |
| ≥ 5 | 67 (0.8) | 58 (0.8) | 9 (1.1) |  |  |
| **Medications** |  |  |  |  |  |
| RAS inhibitor | 1498 (17.9) | 1357 (18.0) | 141 (17.4) | 0.02 | 0.700 |
| Beta-blocker | 2733 (32.7) | 2443 (32.4) | 290 (35.8) | 0.07 | 0.058 |
| MRA | 840 (10.1) | 715 (9.5) | 125 (15.4) | 0.18 | <0.001 |
| Loop diuretics | 2139 (25.6) | 1824 (24.2) | 315 (38.8) | 0.32 | <0.001 |
| Statin | 2660 (31.9) | 2390 (31.7) | 270 (33.3) | 0.03 | 0.377 |
| **Echocardiographic parameters** |  |  |  |  |  |
| LVEF, % | 61.8 (57.8–65.4) | 61.8 (57.8–65.4) | 61.5 (56.7–65.5) | 0.10 | 0.170 |
| LVEDD, mm | 45.0 (42.0–49.0) | 45.0 (42.0–49.0) | 44.0 (40.0–49.0) | 0.15 | <0.001 |
| LVEDS, mm | 29.0 (25.0–33.0) | 29.0 (25.0–33.0) | 28.0 (25.0–33.0) | 0.00 | 0.045 |
| LV mass index, g/m<sup>2</sup> | 87.5 (73.9–103.7) | 87.6 (74.2–103.6) | 84.8 (70.2–104.9) | 0.02 | 0.015 |
| E/e' | 10.1 (8.0–13.2) | 10.0 (7.9–13.0) | 10.9 (8.3–14.7) | 0.25 | <0.001 |
| e', cm/s | 6.4 (5.1–8.0) | 6.4 (5.1–8.0) | 6.0 (4.6–7.4) | 0.19 | <0.001 |
| LA volume index, mL/m<sup>2</sup> | 33.2 (26.5–43.5) | 33.3 (26.6–43.1) | 32.5 (24.4–48.4) | 0.11 | 0.306 |
| TR Vmax, m/s | 2.3 (2.0–2.5) | 2.3 (2.0–2.5) | 2.3 (2.0–2.6) | 0.14 | 0.002 |
| RV FAC, % | 33.1 (26.7–39.1) | 33.3 (26.9–39.2) | 30.4 (21.4–38.0) | 0.19 | <0.001 |

Values are presented as n (%) or median (IQR). P values were calculated using the Mann–Whitney U test for continuous variables and the chi-square test for categorical variables; standardized mean differences (SMDs) are also reported. The modified Charlson comorbidity index (mCCI) was calculated without age adjustment or severity-dependent weighting, assigning 1 point each for myocardial infarction, heart failure, stroke, COPD, liver cirrhosis, and diabetes mellitus, and 2 points each for ESRD and malignancy.
Abbreviations: COPD, chronic obstructive pulmonary disease; ESRD, end-stage renal disease; LA, left atrial; LVEDD, left ventricular end-diastolic diameter; LVEF, left ventricular ejection fraction; LVEDS, left ventricular end-systolic diameter; mCCI, modified Charlson comorbidity index; MRA, mineralocorticoid receptor antagonist; RAS, renin–angiotensin system; RV FAC, right ventricular fractional area change; SMD, standardized mean difference; TR Vmax, peak tricuspid regurgitation velocity.

### Distribution and Echocardiographic Correlates of Multi-Chamber Strain

The median LVGLS, LASr, and RVFWS were 15.2% (IQR, 13.1–17.0), 23.0% (IQR, 17.4–28.4), and 16.2% (IQR, 10.9–20.4), respectively. The distributions of the three strain parameters across clinically relevant subgroups are shown in **Figure 2**. All three strain measures decreased with lower LVEF, with the most pronounced reduction observed for LVGLS. LASr progressively decreased with advancing age, greater LA enlargement, and higher E/e′, whereas RVFWS was substantially lower in patients with reduced RV FAC and decreased with advancing age. LVGLS was lower in men than in women, whereas LASr and RVFWS were broadly similar between sexes.

**Figure 2.**
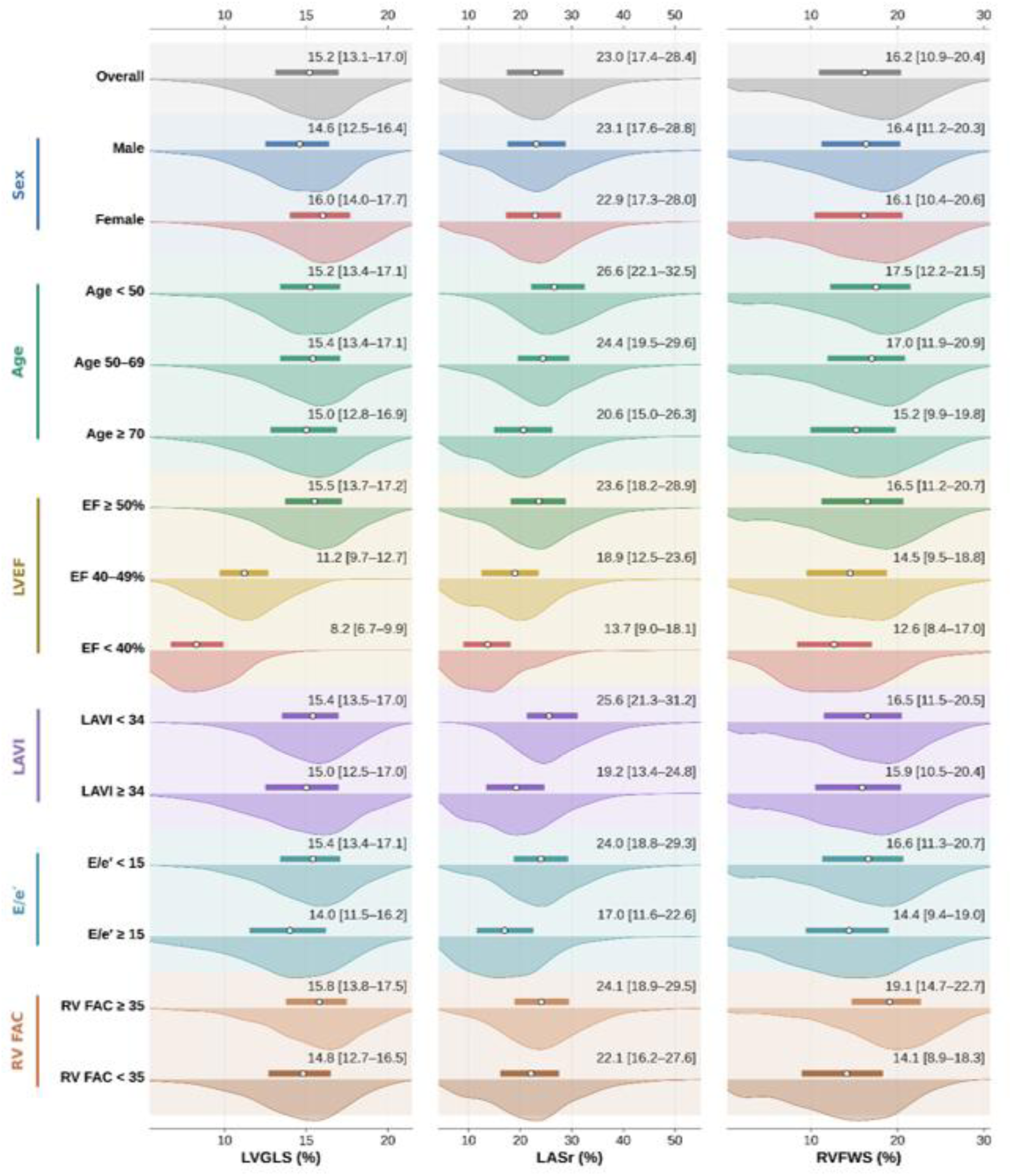
Distribution of Multi-Chamber Strain Across Relevant Subgroups. Distributions of absolute LVGLS, LASr, and RVFWS in the overall cohort and across subgroups defined by sex, age, LVEF, LAVI, E/e′, and RV FAC. Points and bars indicate medians and interquartile ranges.

The AI-derived strain measures showed expected chamber-specific associations with conventional echocardiographic parameters (**Figure 3**). LVGLS correlated moderately with LVEF (Spearman ρ=0.52, P<0.001), LASr was inversely correlated with LAVI (ρ=−0.48, P<0.001), and RVFWS correlated with RV FAC (ρ=0.44, P<0.001). Associations with other structural and hemodynamic parameters were generally weaker.

**Figure 3.**
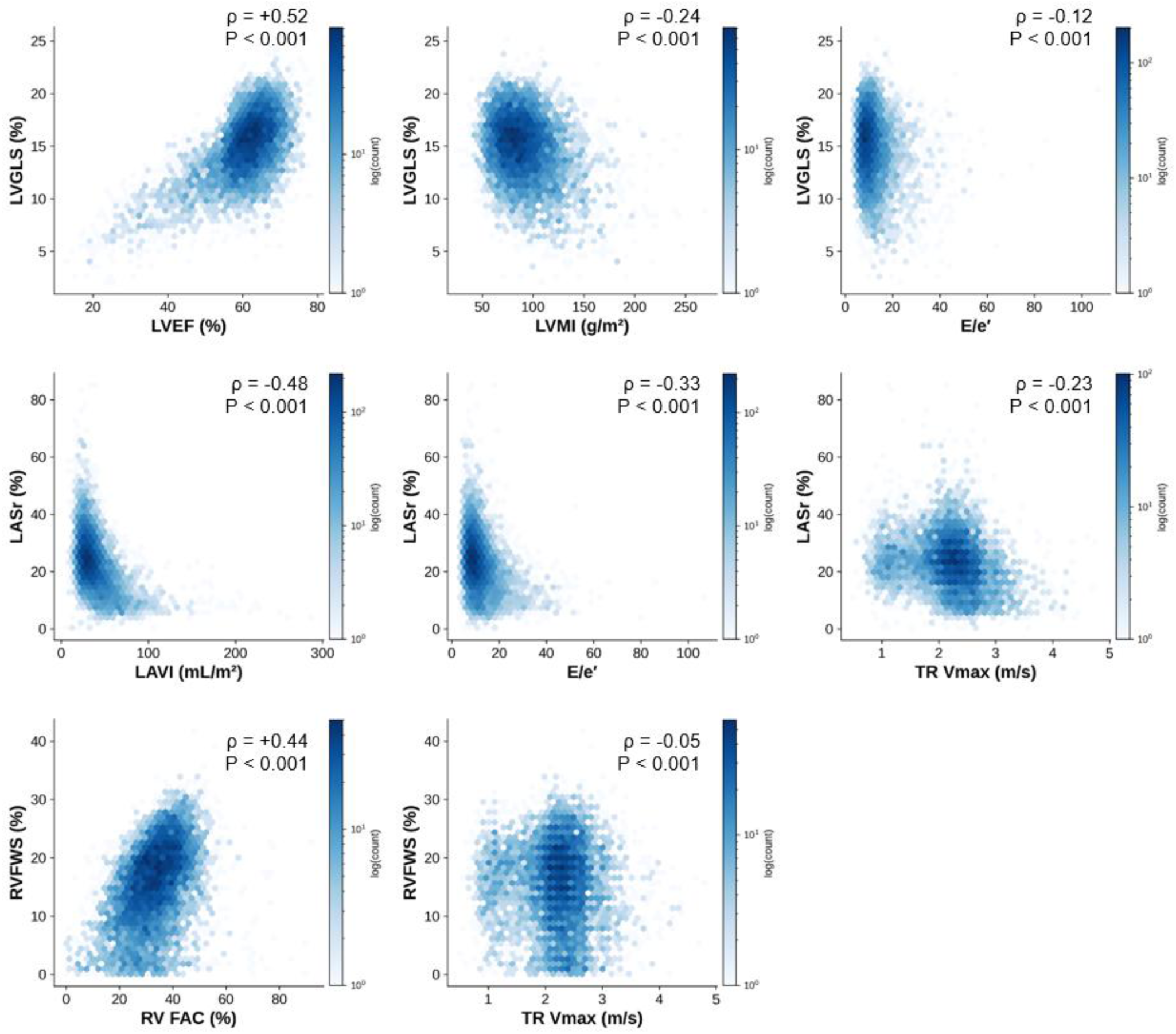
Correlations Between Multi-Chamber Strain and Conventional Echocardiographic Parameters. Hexbin density plots showing associations of LVGLS, LASr, and RVFWS with conventional echocardiographic parameters. Color intensity represents observation density on a logarithmic scale. Spearman correlation coefficients (ρ) and corresponding P values are shown in each panel.

### Associations of Individual Strain Parameters with Clinical Outcomes

During follow-up (median, 6.7 months; IQR, 5.6−8.1), 157 composite outcome events occurred, including 103 all-cause deaths and 60 HF hospitalizations. Restricted cubic spline analyses showed graded associations between lower strain parameters and adverse clinical outcomes, although association strength differed across chambers and outcomes (**Supplemental Result 1**). Lower LVGLS was associated with higher risks of the composite outcome, all-cause death, and HF hospitalization (overall P<0.001, P=0.001, and P<0.001, respectively). Lower LASr was associated with the composite outcome and HF hospitalization (both overall P<0.001), whereas its association with all-cause death was not statistically significant (overall P=0.089). Lower RVFWS was associated with the composite outcome, all-cause death, and HF hospitalization (overall P<0.001, P=0.021, and P<0.001, respectively). These findings were generally consistent in multivariable Cox proportional hazards models evaluating each 5-percentage-point decrease in LVGLS, LASr, and RVFWS (**Supplemental Result 2**).

### Multi-Chamber Strain Clusters and Clinical Outcomes

Based on AI-derived multi-chamber strain measures, three distinct clusters were identified using GMM (**Supplemental Result 3**). The three-cluster solution was selected considering cluster stability, parsimony, and clinical interpretability. The multi-chamber strain profiles of each cluster are visualized in **Figure 4**, with clinical and echocardiographic characteristics summarized in **Table 3** and **Supplemental Result 4**. Cluster 1 was characterized by preserved multi-chamber strain and relatively favorable clinical and echocardiographic profiles. Cluster 2 showed selective RV-predominant dysfunction, with markedly reduced RVFWS despite preserved LVGLS and LASr, and included a higher proportion of women. Cluster 3 exhibited pan-chamber strain impairment and was characterized by older age, male predominance, greater comorbidity burden, and more adverse conventional echocardiographic features.

**Figure 4.**
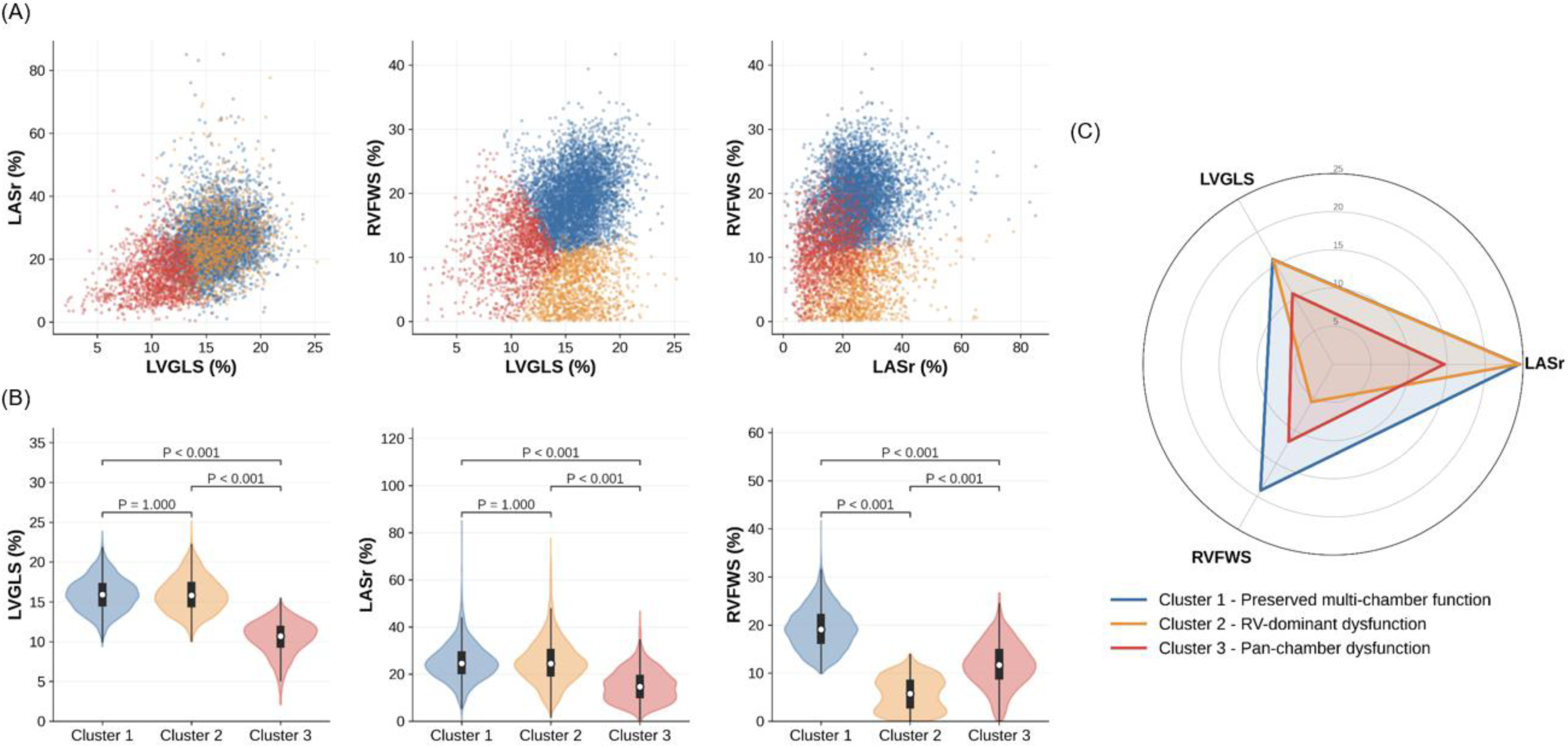
Identification and Multi-Chamber Strain Profiles of the Three Clusters. Gaussian mixture model clustering based on standardized absolute LVGLS, LASr, and RVFWS. (A) Pairwise distributions of the strain measures by cluster. (B) Violin plots showing cluster-specific distributions, with medians and interquartile ranges; pairwise comparisons used the Mann– Whitney U test with Bonferroni correction. (C) Radar plot showing the median multi-chamber strain profile of each cluster.

**Table 3.** Clinical Characteristics and Outcomes According to Cluster Assignment.

| **Variable** | **Overall**<br>**(N = 7,539)** | **Cluster 1**<br>***Preserved***<br>**(N = 4,796)** | **Cluster 2**<br>***RV-dominant***<br>**(N = 1,324)** | **Cluster 3**<br>***Pan-chamber***<br>**(N = 1,419)** | **P value** |
| --- | --- | --- | --- | --- | --- |
| **Demographics** |  |  |  |  |  |
| Age, years | 69.0 (60.0–77.0) | 68.0 (59.0–76.0) | 70.0 (60.0–77.0) | 72.0 (62.0–80.0) | <0.001 |
| Male | 4366 (57.9) | 2733 (57.0) | 652 (49.2) | 981 (69.1) | <0.001 |
| **Comorbidities** |  |  |  |  |  |
| Hypertension | 1678 (22.3) | 1012 (21.1) | 271 (20.5) | 395 (27.8) | <0.001 |
| Diabetes mellitus | 392 (5.2) | 234 (4.9) | 51 (3.9) | 107 (7.5) | <0.001 |
| Heart failure | 1102 (14.6) | 478 (10.0) | 138 (10.4) | 486 (34.2) | <0.001 |
| Myocardial infarction | 605 (8.0) | 323 (6.7) | 82 (6.2) | 200 (14.1) | <0.001 |
| Atrial fibrillation | 1211 (16.1) | 525 (10.9) | 206 (15.6) | 480 (33.8) | <0.001 |
| COPD | 211 (2.8) | 111 (2.3) | 36 (2.7) | 64 (4.5) | <0.001 |
| Stroke | 881 (11.7) | 508 (10.6) | 149 (11.3) | 224 (15.8) | <0.001 |
| ESRD | 215 (2.9) | 130 (2.7) | 18 (1.4) | 67 (4.7) | <0.001 |
| Liver cirrhosis | 98 (1.3) | 61 (1.3) | 20 (1.5) | 17 (1.2) | 0.740 |
| Malignancy | 1659 (22.0) | 1034 (21.6) | 331 (25.0) | 294 (20.7) | 0.012 |
| mCCI | 1.0 (0.0–2.0) | 0.0 (0.0–2.0) | 0.0 (0.0–2.0) | 1.0 (0.0–2.0) | <0.001 |
| 0 | 3742 (49.6) | 2597 (54.1) | 681 (51.4) | 464 (32.7) |  |
| 1–2 | 3072 (40.7) | 1818 (37.9) | 532 (40.2) | 722 (50.9) |  |
| 3–4 | 667 (8.8) | 348 (7.3) | 107 (8.1) | 212 (14.9) |  |
| ≥ 5 | 58 (0.8) | 33 (0.7) | 4 (0.3) | 21 (1.5) |  |
| **Medications** |  |  |  |  |  |
| RAS inhibitor | 1357 (18.0) | 705 (14.7) | 167 (12.6) | 485 (34.2) | <0.001 |
| Beta-blocker | 2443 (32.4) | 1345 (28.0) | 349 (26.4) | 749 (52.8) | <0.001 |
| MRA | 715 (9.5) | 287 (6.0) | 82 (6.2) | 346 (24.4) | <0.001 |
| Loop diuretics | 1824 (24.2) | 925 (19.3) | 277 (20.9) | 622 (43.8) | <0.001 |
| Statin | 2390 (31.7) | 1436 (29.9) | 374 (28.2) | 580 (40.9) | <0.001 |
| **Echocardiographic parameters** |  |  |  |  |  |
| LVEF, % | 61.8 (57.8–65.4) | 62.6 (59.2–65.8) | 62.7 (59.6–66.0) | 55.3 (44.4–61.0) | <0.001 |
| LVEDD, mm | 45.0 (42.0–49.0) | 45.0 (42.0–48.0) | 45.0 (41.0–48.0) | 47.0 (43.0–52.0) | <0.001 |
| LVEDS, mm | 29.0 (25.0–33.0) | 28.0 (25.0–32.0) | 28.0 (25.0–31.0) | 33.0 (28.0–39.0) | <0.001 |
| LV mass index, g/m<sup>2</sup> | 87.6 (74.2–103.6) | 86.3 (73.3–100.9) | 83.7 (71.0–97.7) | 99.1 (82.1–120.5) | <0.001 |
| E/e' | 10.0 (7.9–13.0) | 9.7 (7.8–12.3) | 10.0 (7.8–12.7) | 11.6 (8.7–16.3) | <0.001 |
| e', cm/s | 6.4 (5.1–8.0) | 6.6 (5.3–8.1) | 6.6 (5.2–8.4) | 5.5 (4.3–7.2) | <0.001 |
| LA volume index, mL/m<sup>2</sup> | 33.3 (26.6–43.1) | 32.4 (26.5–40.4) | 31.7 (25.8–41.2) | 40.7 (29.4–56.6) | <0.001 |
| TR Vmax, m/s | 2.3 (2.0–2.5) | 2.2 (2.0–2.5) | 2.3 (2.0–2.5) | 2.3 (2.0–2.6) | <0.001 |
| RV FAC, % | 33.5 (27.3–39.3) | 35.3 (30.1–40.6) | 28.6 (21.8–35.5) | 29.2 (23.5–35.5) | <0.001 |
| **Multi-chamber strain** |  |  |  |  |  |
| LVGLS, % | 15.2 (13.1–17.0) | 15.9 (14.4–17.4) | 15.8 (14.3–17.5) | 10.7 (9.2–12.0) | <0.001 |
| LASr, % | 23.0 (17.4–28.4) | 24.5 (19.9–29.6) | 24.4 (19.0–30.6) | 14.6 (9.8–19.8) | <0.001 |
| RVFWS, % | 16.2 (10.9–20.4) | 19.1 (16.1–22.3) | 5.7 (2.6–8.6) | 11.7 (8.6–15.0) | <0.001 |

Values are presented as n (%) or median (IQR). P values were calculated using the Kruskal–Wallis test for continuous variables and the chi-square test for categorical variables.
Abbreviations: COPD, chronic obstructive pulmonary disease; ESRD, end-stage renal disease; HF, heart failure; LASr, left atrial reservoir strain; LVEDD, left ventricular end-diastolic diameter; LVEF, left ventricular ejection fraction; LVEDS, left ventricular end-systolic diameter; LVGLS, left ventricular global longitudinal strain; mCCI, modified Charlson comorbidity index; MRA, mineralocorticoid receptor antagonist; RAS, renin–angiotensin system; RV FAC, right ventricular fractional area change; RVFWS, right ventricular free-wall strain; TR Vmax, peak tricuspid regurgitation velocity.

In multivariable analyses, Cluster 3 was consistently associated with worse clinical outcomes compared with Cluster 1 (**Figure 5**). Cluster 3 was associated with the highest risk of the composite outcome (adjusted HR, 3.44; 95% CI, 2.43–4.88; P<0.001), all-cause death (adjusted HR, 2.11; 95% CI, 1.36–3.29; P<0.001), and HF hospitalization (adjusted HR, 7.60; 95% CI, 4.18–13.82; P<0.001) (**Supplemental Result 5**). Cluster 2 was not significantly associated with the composite outcome (adjusted HR, 1.22; 95% CI, 0.74–2.01; P=0.437) or all-cause death (adjusted HR, 1.64; 95% CI, 0.98–2.77; P=0.061); no HF hospitalization events occurred in Cluster 2, precluding estimation of the corresponding hazard ratio. Addition of cluster assignment in Model 2 significantly improved overall model fit compared with Model 1 for all three outcomes (**Supplemental Result 6**). Among patients with LVEF ≥50%, the pattern of associations was generally consistent with that observed in the full cohort (**Supplemental Result 7**). Cluster 3 remained significantly associated with all three outcomes, whereas Cluster 2 was associated only with all-cause death (adjusted HR, 1.73; 95% CI, 1.02–2.92; P=0.041).

**Figure 5.**
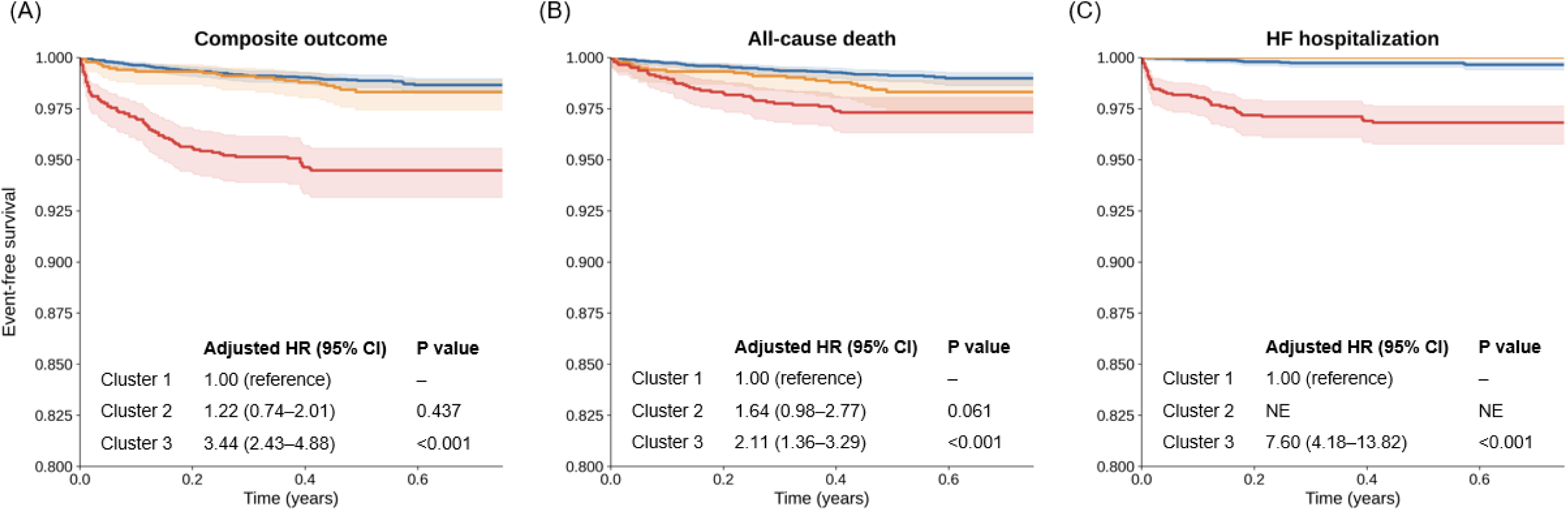
Kaplan–Meier Curves for Clinical Outcomes According to Cluster Assignment. Kaplan–Meier curves for (A) the composite outcome, (B) all-cause death, and (C) HF hospitalization by cluster. The hazard ratio for Cluster 2 was not estimable for HF hospitalization because no events occurred during follow-up period. Abbreviation: NE, not estimable.

## DISCUSSION

In this study of consecutive patients undergoing routine TTE, complete and fully automated assessment of LV, LA, and RV strain was feasible in 90.3% of examinations without manual intervention. The three strain measures showed different patterns across clinical subgroups, chamber-specific relationships with conventional echocardiographic parameters, and differential associations with clinical outcomes. Integration of LVGLS, LASr, and RVFWS identified three distinct clusters characterized by preserved multi-chamber function, RV-dominant dysfunction, and pan-chamber dysfunction. Among these, the pan-chamber dysfunction cluster was associated with higher risks of all-cause death and HF hospitalization. The addition of cluster assignment improved model fit beyond clinical variables, LVEF, and RV FAC, and similar prognostic associations were observed among patients with LVEF ≥50%. These findings suggest that patterns of dysfunction across cardiac chambers may provide complementary prognostic information beyond conventional echocardiographic measures.

### Feasibility of Automated Multi-Chamber Strain Assessment in Routine Practice

Despite extensive evidence supporting myocardial strain, its routine use remains limited by the need for view selection, myocardial contouring, tracking verification, and manual correction, while differences in post-processing algorithms may introduce additional variability.^3^ Extending strain assessment beyond LVGLS to LA and RV further increases this complexity.^3^ In the present study, however, fully automated analysis provided complete multi-chamber strain measurements in approximately 90% of consecutive TTE examinations without additional image acquisition or manual processing. These findings extend prior work on automated echocardiographic analysis by demonstrating the feasibility of hospital-wide multi-chamber strain assessment in routine clinical practice.

### Complementary Functional Information Across Cardiac Chambers

LVGLS, LASr, and RVFWS each showed expected associations with conventional chamber-specific measures, including LVEF, LAVI, and RV FAC, respectively; however, these correlations were only modest, suggesting that strain captures complementary rather than interchangeable aspects of cardiac function. LVGLS reflects longitudinal myocardial shortening beyond volumetric LVEF, LASr integrates atrial myocardial function and loading conditions, and RVFWS directly assesses RV myocardial deformation despite the geometric limitations of conventional RV assessment.^3,6,14–17^ Previous studies have also demonstrated incremental prognostic value of LVGLS beyond LVEF, LASr in relation to atrial function and HF outcomes, and RVFWS beyond conventional indices of RV function.^6,14–17^ The three strain measures also showed distinct patterns across clinical subgroups and differential associations with clinical outcomes. These findings support a multi-chamber approach in which LV, LA, and RV strain provide complementary information on overall cardiac functional burden.

### Multi-Chamber Strain Clusters and Clinical Risk

A major finding of this study was that integration of only three chamber-specific deformation measures identified distinct patterns of multi-chamber dysfunction with different clinical profiles and outcomes. This extends previous applications of data-driven clustering in heterogeneous cardiovascular populations by demonstrating that a small set of strain measures can capture clinically relevant heterogeneity in cardiac functional involvement.^18–21^

The pan-chamber dysfunction cluster was associated with the highest risk of adverse clinical outcomes, particularly HF hospitalization, suggesting that impairment across multiple chambers may reflect a greater overall burden of cardiac dysfunction than predominantly single-chamber involvement. In contrast, the RV-dominant cluster showed markedly reduced RVFWS despite relatively preserved LVGLS and LASr, indicating a distinct pattern rather than generalized cardiac dysfunction. Although this cluster had distinct clinical characteristics, including a higher prevalence of malignancy, its underlying mechanisms and clinical significance remain uncertain and warrant further investigation.

### Limitations

Several limitations should be acknowledged. First, this retrospective, single-center study may limit the generalizability to other institutions, populations, imaging protocols and ultrasound systems. Nevertheless, consecutive enrollment across diverse clinical settings provides a relatively unselected population. Second, the short follow-up resulted in a limited number of events for some outcomes, particularly within the RV-dominant cluster; therefore, subgroup-specific associations should be interpreted cautiously, and longer follow-up is needed to better define their prognostic significance. Third, multi-chamber strain analysis was not feasible in all examinations. Unsuccessful analysis was more common in patients with heart failure, atrial fibrillation, and lower RV FAC, raising the possibility of selection bias and indicating nonuniform performance across clinical or image-quality conditions. Fourth, residual confounding cannot be excluded despite multivariable adjustment, and supplementation of missing conventional echocardiographic measurements with AI-derived values may have introduced measurement heterogeneity. Finally, the cluster assignment may depend on the study population, analytic approach, and AI platform, warranting external validation in independent cohorts.

## Conclusions

Fully automated, simultaneous multi-chamber strain assessment was feasible in a large consecutive cohort undergoing routine TTE. Integration of these chamber-specific strain measures identified distinct patterns of cardiac functional involvement, with pan-chamber dysfunction cluster associated with the highest clinical risk. These findings support automated multi-chamber strain profiling as a scalable framework for comprehensive functional characterization in routine echocardiographic practice and provide a basis for future external and prospective validation.

## Data Availability

The data underlying this study are not publicly available due to ethical restrictions and patient privacy considerations. De-identified data may be made available from the corresponding author upon reasonable request.

## Author contributoions

Su A Noh (Data curation [equal], Methodology [lead], Writing—original draft [lead]), Jiesuck Park (Data curation, Conceptualization, Writing—review & editing [equal]), Yeonyee E. Yoon (Conceptualization, Writing—original draft [lead], Writing—review & editing [lead]), Jiyeon Kim (Writing—review & editing [supporting]), Jaeik Jeon (Writing—review & editing [supporting]), Yeonggul Jang (Writing—review & editing [supporting]), Seung-Ah Lee (Writing—review & editing [supporting]), Minjung Bak (Writing—review & editing [supporting]), Hong-Mi Choi (Writing—review & editing [supporting]), In-Chang Hwang (Writing—review & editing [supporting]), Goo-Yeong Cho (Writing—review & editing [supporting])

## Data availability

The data underlying this study cannot be made publicly available due to ethical restrictions set by the IRB of the study institution; i.e., public availability would compromise patient confidentiality and participant privacy. Please contact the corresponding authors to request the minimal anonymized dataset. Researchers with additional inquiries about the deep learning model developed in this study are also encouraged to reach out to the corresponding authors.

## Supplemental Methods

## Supplemental Method 1. Study Flow and Overview of Multi-Chamber Strain Analyses

The feasibility cohort comprised 8,350 consecutive patients who underwent transthoracic echocardiography at Seoul National University Bundang Hospital between August and December 2025 and in whom fully automated strain analysis was attempted as part of the routine workflow. Feasibility was assessed in the entire cohort and was defined by the successful acquisition of AI-derived strain measurements in the corresponding cardiac chambers. Multi-chamber strain analysis was defined as the simultaneous availability of 3-view left ventricular global longitudinal strain (LVGLS), apical 4-chamber (A4C)-derived left atrial reservoir strain (LASr), and right ventricular free-wall strain (RVFWS). Multi-chamber strain analysis was successfully obtained in 7,539 patients (90.3%), who constituted the study population for subsequent analysis. In 811 patients (9.7%), multi-chamber strain analysis was unsuccessful because at least one required chamber-specific strain measurement was unavailable.

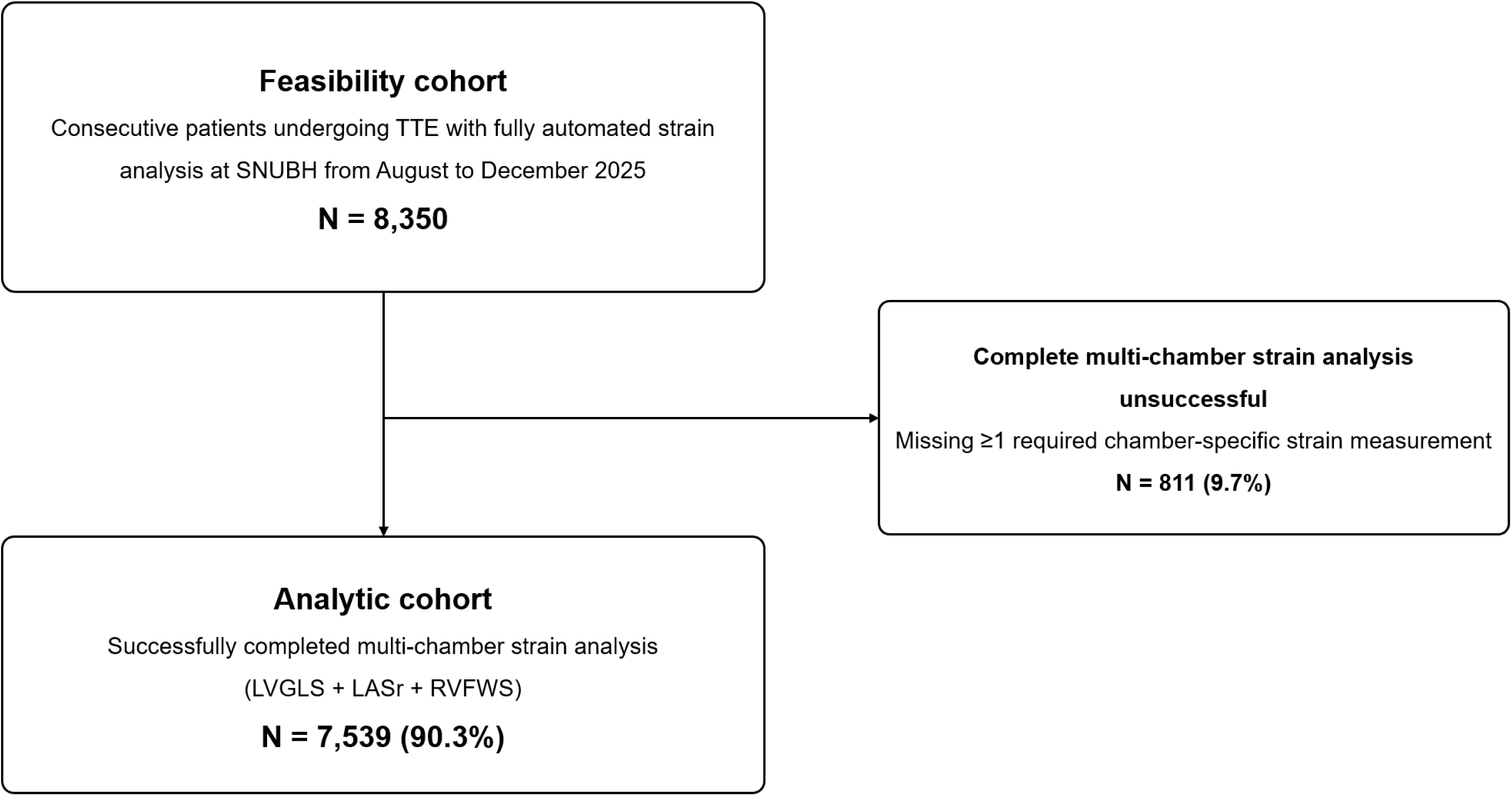

Abbreviations: A4C, apical 4-chamber; AI, artificial intelligence; LASr, left atrial reservoir strain; LVGLS, left ventricular global longitudinal strain; RVFWS, right ventricular free-wall strain; TTE, transthoracic echocardiography.

## Supplemental Method 2. Fully Automated Multi-Chamber Strain Analysis

Within the automated analysis workflow described above, image loops for strain analysis were selected according to prespecified chamber-specific view priorities and segmentation-based quality scores.^1–5^ When multiple cine loops were available within the same view category, the cine loop with the most favorable quality score was selected. LVGLS, LASr, and RVFWS were generated through automated myocardial motion tracking without manual view selection, contour adjustment, tracking correction, or parameter entry.

For LV strain analysis, zoomed apical views were prioritized when available. LVGLS was calculated as the average of valid longitudinal strain measurements obtained from the apical 4-chamber, 2-chamber, and 3-chamber views (A4C, A2C, and A3C, respectively). A complete three-view LVGLS measurement required successful analysis of all three apical views. For LA strain analysis, LASr was derived from the A4C in accordance with current consensus recommendations.^6^ A2C-derived LASr was assessed only for view-specific technical feasibility and was not used in the primary analysis or to define successful multi-chamber strain analysis. For RV strain analysis, an RV-focused apical view was prioritized over a standard A4C view for RVFWS calculation.

A chamber-specific strain measurement was considered feasible when the system generated a valid strain value after automated view selection, segmentation, and motion tracking. When no suitable view was available or predefined requirements for segmentation, measurement, or motion tracking were not met, the corresponding parameter was not generated. Unsuccessful analyses were not manually corrected or retrospectively reanalyzed.

## Supplemental Method 3. Availability and AI Substitution of Conventional Echocardiographic Measurements in the Analytic Cohort (N=7,539)

Conventional echocardiographic parameters were preferentially obtained from manual measurements, with AI-derived values substituted when manual measurements were unavailable.

| Parameter | N with final value | Conventional measurement available, N (%) | AI-derived measurement substituted, N (%) | Conventional measurement only, median (IQR) | After AI substitution, median (IQR) |
| --- | --- | --- | --- | --- | --- |
| LVEF, % | 7539 | 7530 (99.9) | 9 (0.1) | 61.8 (57.8–65.4) | 61.8 (57.8–65.4) |
| LVEDD, mm | 7537 | 7531 (99.9) | 6 (0.1) | 45.0 (42.0–49.0) | 45.0 (42.0–49.0) |
| LVESD, mm | 7538 | 7530 (99.9) | 8 (0.1) | 29.0 (25.0–33.0) | 29.0 (25.0–33.0) |
| LV mass index, g/m<sup>2</sup> | 7536 | 7529 (99.9) | 7 (0.1) | 87.6 (74.2–103.6) | 87.6 (74.2–103.6) |
| LA volume index, mL/m<sup>2</sup> | 7528 | 7473 (99.1) | 55 (0.7) | 33.3 (26.7–43.2) | 33.3 (26.6–43.1) |
| E/e' | 7529 | 7221 (95.8) | 308 (4.1) | 10.0 (7.9–12.9) | 10.0 (7.9–13.0) |
| e', cm/s | 7535 | 7236 (96.0) | 299 (4.0) | 6.4 (5.1–8.0) | 6.4 (5.1–8.0) |
| TR Vmax, m/s | 7492 | 5772 (76.6) | 1720 (22.8) | 2.3 (2.2–2.6) | 2.3 (2.0–2.5) |
| RV FAC, % | 7507 | 292 (3.9) | 7215 (95.7) | 35.0 (28.0–38.0) | 33.5 (27.3–39.3) |

Categorical variables are presented as n (%), and continuous variables as median (IQR).
Abbreviations: AI, artificial intelligence; IQR, interquartile range; LA, left atrial; LV, left ventricular; LVEDD, left ventricular end-diastolic diameter; LVEF, left ventricular ejection fraction; LVESD, left ventricular end-systolic diameter; RV FAC, right ventricular fractional area change; TR Vmax, peak tricuspid regurgitation velocity.

## Supplemental Method 4. Agreement Between Manual and AI-derived Measurements in Patients with Both Measurements Available

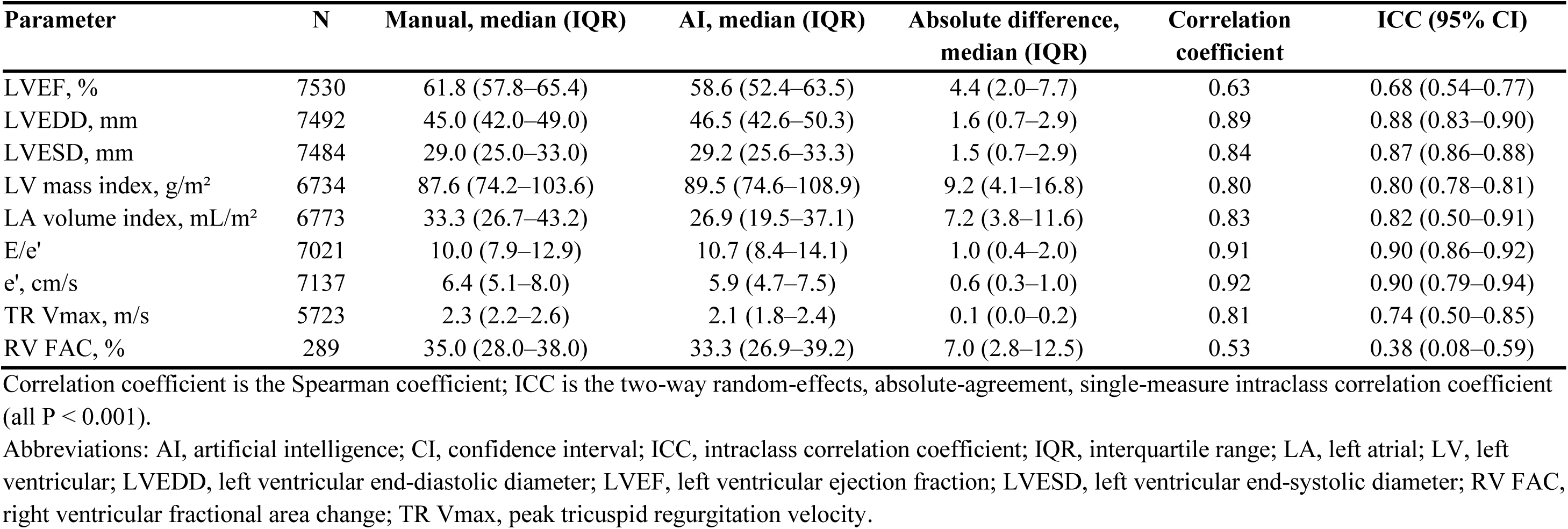

| Parameter | N | Manual, median (IQR) | AI, median (IQR) | Absolute difference, median (IQR) | Correlation coefficient | ICC (95% CI) |
| --- | --- | --- | --- | --- | --- | --- |
| LVEF, % | 7530 | 61.8 (57.8–65.4) | 58.6 (52.4–63.5) | 4.4 (2.0–7.7) | 0.63 | 0.68 (0.54–0.77) |
| LVEDD, mm | 7492 | 45.0 (42.0–49.0) | 46.5 (42.6–50.3) | 1.6 (0.7–2.9) | 0.89 | 0.88 (0.83–0.90) |
| LVESD, mm | 7484 | 29.0 (25.0–33.0) | 29.2 (25.6–33.3) | 1.5 (0.7–2.9) | 0.84 | 0.87 (0.86–0.88) |
| LV mass index, g/m<sup>2</sup> | 6734 | 87.6 (74.2–103.6) | 89.5 (74.6–108.9) | 9.2 (4.1–16.8) | 0.80 | 0.80 (0.78–0.81) |
| LA volume index, mL/m<sup>2</sup> | 6773 | 33.3 (26.7–43.2) | 26.9 (19.5–37.1) | 7.2 (3.8–11.6) | 0.83 | 0.82 (0.50–0.91) |
| E/e' | 7021 | 10.0 (7.9–12.9) | 10.7 (8.4–14.1) | 1.0 (0.4–2.0) | 0.91 | 0.90 (0.86–0.92) |
| e', cm/s | 7137 | 6.4 (5.1–8.0) | 5.9 (4.7–7.5) | 0.6 (0.3–1.0) | 0.92 | 0.90 (0.79–0.94) |
| TR Vmax, m/s | 5723 | 2.3 (2.2–2.6) | 2.1 (1.8–2.4) | 0.1 (0.0–0.2) | 0.81 | 0.74 (0.50–0.85) |
| RV FAC, % | 289 | 35.0 (28.0–38.0) | 33.3 (26.9–39.2) | 7.0 (2.8–12.5) | 0.53 | 0.38 (0.08–0.59) |

Correlation coefficient is the Spearman coefficient; ICC is the two-way random-effects, absolute-agreement, single-measure intraclass correlation coefficient (all $P < 0.001$ ).
Abbreviations: AI, artificial intelligence; CI, confidence interval; ICC, intraclass correlation coefficient; IQR, interquartile range; LA, left atrial; LV, left ventricular; LVEDD, left ventricular end-diastolic diameter; LVEF, left ventricular ejection fraction; LVESD, left ventricular end-systolic diameter; RV FAC, right ventricular fractional area change; TR Vmax, peak tricuspid regurgitation velocity.

## Supplemental Results

## Supplemental Result 1. Restricted Cubic Spline Analysis of Multi-Chamber Strain and Clinical Outcomes

Adjusted hazard ratios for the composite outcome, all-cause death, and HF hospitalization across the continuous range of LVGLS, LASr, and RVFWS. Solid lines indicate hazard ratios and shaded areas indicate 95% confidence intervals, dotted horizontal lines indicate a hazard ratio (HR) of 1, and vertical lines indicate cohort medians used as the reference values. Models were adjusted for age, sex, and the modified Charlson comorbidity index, with 3 knots at the 10^th^, 50^th^, and 90^th^ percentiles.

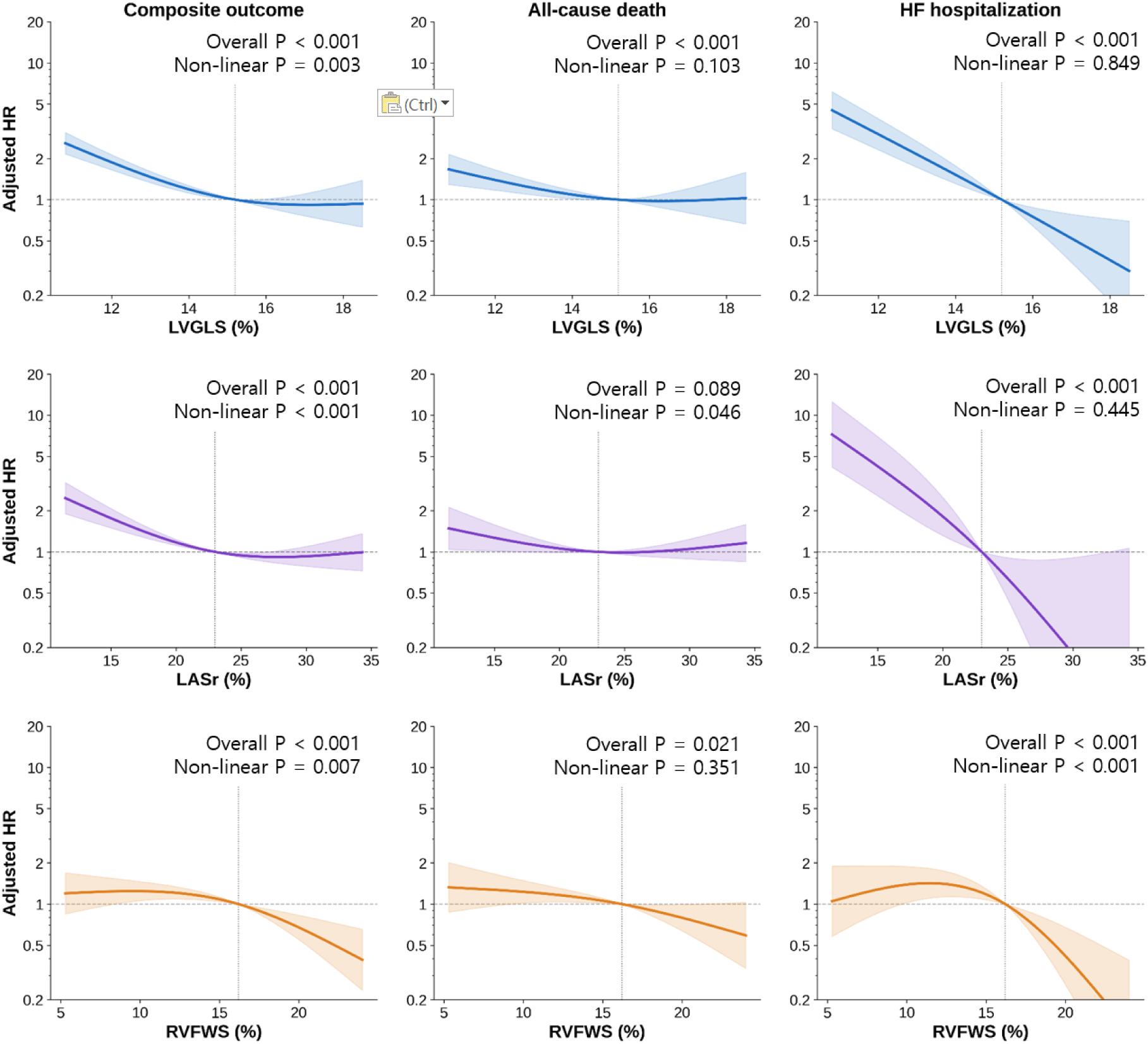

Abbreviations: HF, heart failure; HR, hazard ratio; LASr, left atrial reservoir strain; LVGLS, left ventricular global longitudinal strain; RVFWS, right ventricular free-wall strain.

## Supplemental Result 2. Multivariable Cox Proportional Hazards Models for Clinical Outcomes According To sSrain Parameters

Hazard ratios are presented per 5-percentage-point decrease in each strain parameter. Each strain parameter was evaluated in a separate multivariable Cox proportional hazards model adjusted for age, sex, and modified Charlson comorbidity index.

| Variable | Composite outcome |  | All-cause death |  | HF hospitalization |  |
| --- | --- | --- | --- | --- | --- | --- |
|  | HR (95% CI) | P value | HR (95% CI) | P value | HR (95% CI) | P value |
| LVGLS | 2.79 (2.24–3.49) | <0.001 | 1.63 (1.22–2.17) | <0.001 | 6.20 (4.41–8.72) | <0.001 |
| LASr | 1.30 (1.18–1.44) | <0.001 | 1.05 (0.94–1.18) | 0.362 | 2.29 (1.87–2.81) | <0.001 |
| RVFWS | 1.24 (1.11–1.39) | <0.001 | 1.21 (1.05–1.39) | 0.009 | 1.31 (1.09–1.58) | 0.004 |

Abbreviations: CI, confidence interval; HF, heart failure; HR, hazard ratio; LASr, left atrial reservoir strain; LVGLS, left ventricular global longitudinal strain; RVFWS, right ventricular free-wall strain.

## Supplemental Result 3. Selection of the Number of Clusters

Candidate cluster solutions from k=2 to k=6 were evaluated using the Bayesian information criterion (BIC), integrated completed likelihood (ICL), silhouette coefficient, and bootstrap-adjusted Rand index (ARI). BIC and ICL favored a four-cluster solution, whereas bootstrap stability was highest for three clusters. The silhouette coefficient decreased with increasing cluster number. Considering cluster stability, parsimony, and clinical interpretability, a three-cluster solution was selected for subsequent analyses.

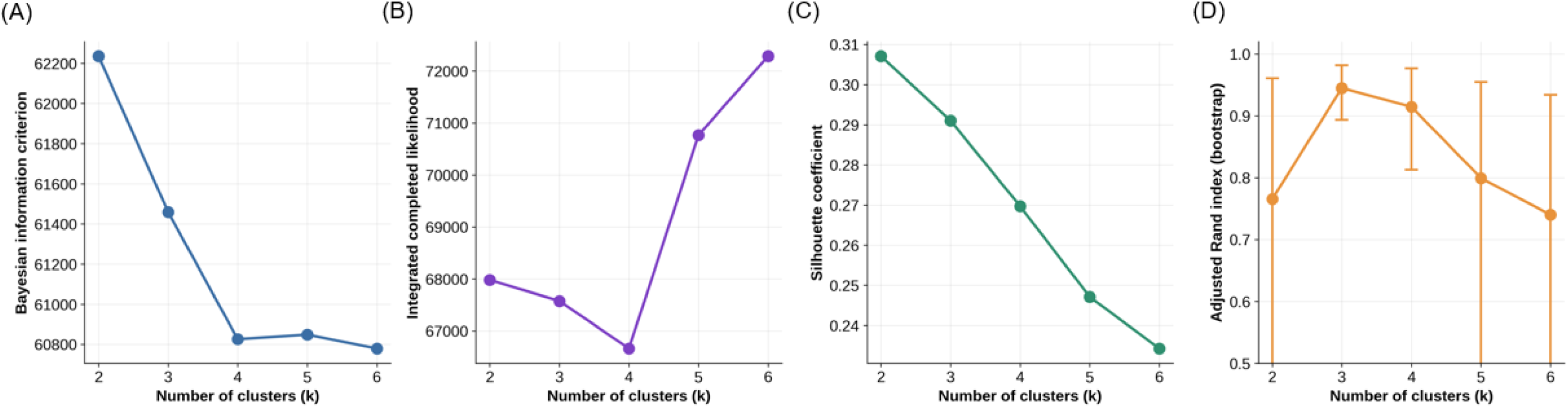

Model-selection metrics used to evaluate candidate cluster solutions (k=2–6): (A) Bayesian information criterion (BIC), (B) integrated completed likelihood (ICL), (C) silhouette coefficient, and (D) bootstrap-adjusted Rand index (ARI).

## Supplemental Result 4. Clinical and Echocardiographic Characteristics Across Clusters

Standardized clinical, echocardiographic, and medication characteristics across the three clusters. Red and blue indicate values above and below the cohort mean, respectively.

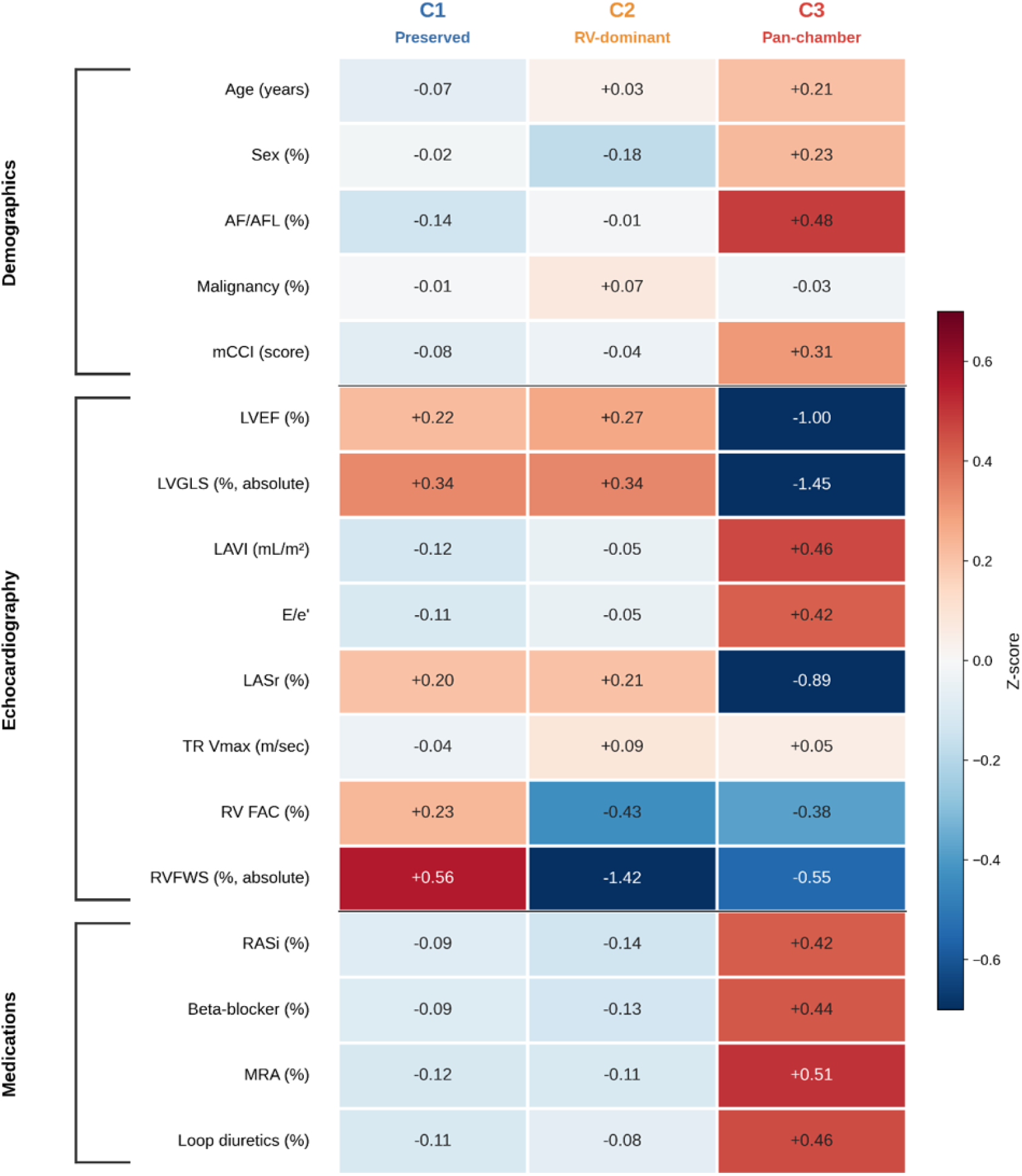

Values are standardized z-scores; color intensity is truncated at ±0.7 SD.

Abbreviations: AF/AFL, atrial fibrillation/flutter; LASr, left atrial reservoir strain; LAVI, left atrial volume index; LVEF, left ventricular ejection fraction; LVGLS, left ventricular global longitudinal strain; mCCI, modified Charlson comorbidity index; MRA, mineralocorticoid receptor antagonist; RASi, renin-angiotensin system inhibitor; RV FAC, right ventricular fractional area change; RVFWS, right ventricular free-wall strain; SD, standard deviation; TR Vmax, peak tricuspid regurgitation velocity.

## Supplemental Result 5. Multivariable Cox Proportional Hazards models for Clinical Outcomes according to Cluster Assignment

Multivariable Cox proportional hazards models were adjusted for age, sex, and modified Charlson comorbidity index (mCCI). Cluster 1 was used as the reference group. Hazard ratios (HRs) for age and mCCI represent per-year and per-point increases, respectively.

| Variable | Composite outcome |  | All-cause death |  | HF hospitalization |  |
| --- | --- | --- | --- | --- | --- | --- |
|  | HR (95% CI) | P value | HR (95% CI) | P value | HR (95% CI) | P value |
| Age, per year | 1.03 (1.02–1.05) | <0.001 | 1.03 (1.01–1.05) | <0.001 | 1.04 (1.02–1.07) | 0.001 |
| Male sex | 0.73 (0.52–1.01) | 0.054 | 0.91 (0.60–1.36) | 0.632 | 0.54 (0.32–0.91) | 0.022 |
| mCCI, per point | 1.60 (1.44–1.78) | <0.001 | 1.59 (1.39–1.81) | <0.001 | 1.59 (1.34–1.88) | <0.001 |
| Cluster Assignment |  |  |  |  |  |  |
| Cluster 1 | 1.00 (reference) | — | 1.00 (reference) | — | 1.00 (reference) | — |
| Cluster 2 | 1.22 (0.74–2.01) | 0.437 | 1.64 (0.98–2.77) | 0.061 | NE | NE |
| Cluster 3 | 3.44 (2.43–4.88) | <0.001 | 2.11 (1.36–3.29) | <0.001 | 7.60 (4.18–13.82) | <0.001 |

Abbreviations: CI, confidence interval; HF, heart failure; HR, hazard ratio; mCCI, modified Charlson comorbidity index; NE, not estimable.

## Supplemental Result 6. Incremental Prognostic Value of Strain Cluster Assignment Beyond Clinical Factors, LVEF and RV FAC

Global likelihood-ratio χ² statistics are shown for the composite outcome (A), all-cause death (B), and HF hospitalization (C). Model 1 was the clinical model including age, sex, mCCI, LVEF, and RV FAC, whereas Model 2 additionally included cluster assignment.

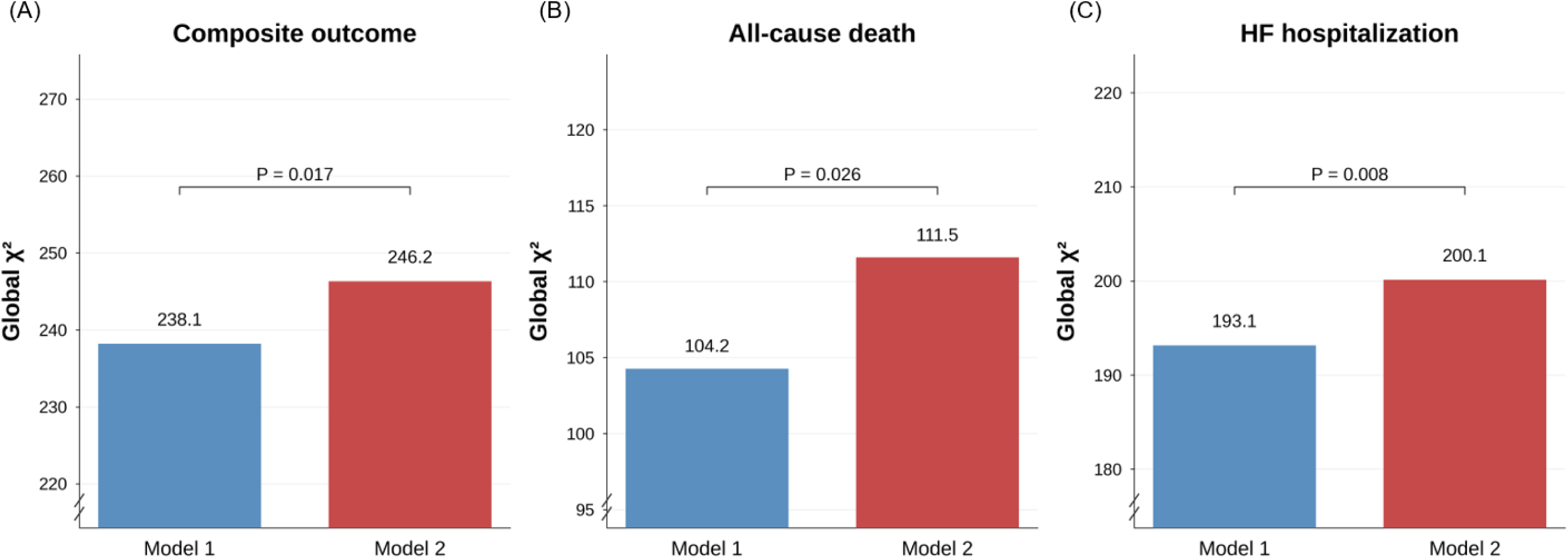

The incremental contribution of cluster assignment was assessed using a likelihood-ratio test comparing Model 2 with Model 1. Brackets indicate likelihood-ratio tests comparing the two models.

Abbreviations: HF, heart failure; LVEF, left ventricular ejection fraction; mCCI, modified Charlson comorbidity index; RV FAC, right ventricular fractional area change.

## Supplemental Result 7. Kaplan–Meier Curves for Clinical Outcomes According to Cluster Assignment in Patients with LVEF ≥50%

Kaplan–Meier curves for (A) the composite outcome, (B) all-cause death, and (C) HF hospitalization according to cluster assignment among patients with LVEF ≥50%.

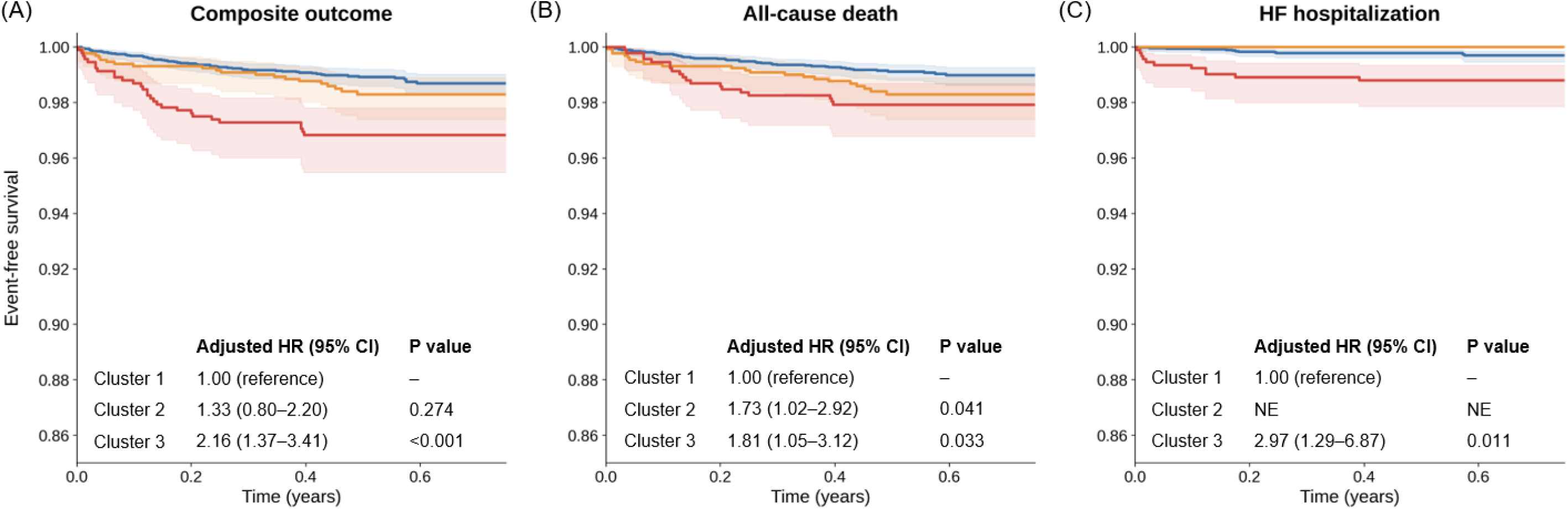

Abbreviations: CI, confidence interval; HF, heart failure; HR, hazard ratio; LVEF, left ventricular ejection fraction.

